# The role of absolute humidity on transmission rates of the COVID-19 outbreak

**DOI:** 10.1101/2020.02.12.20022467

**Authors:** Wei Luo, Maimuna S. Majumder, Diambo Liu, Canelle Poirier, Kenneth D Mandl, Marc Lipsitch, Mauricio Santillana

**Author notes:** Corresponding author: Mauricio Santillana.

## Abstract

A novel coronavirus (COVID-19) was identified in Wuhan, Hubei Province, China, in December 2019 and has caused over 40,000 cases worldwide to date. Previous studies have supported an epidemiological hypothesis that cold and dry (low absolute humidity) environments facilitate the survival and spread of droplet-mediated viral diseases, and warm and humid (high absolute humidity) environments see attenuated viral transmission (i.e., influenza). However, the role of absolute humidity in transmission of COVID-19 has not yet been established. Here, we examine province-level variability of the basic reproductive numbers of COVID-19 across China and find that changes in weather alone (i.e., increase of temperature and humidity as spring and summer months arrive in the North Hemisphere) will not necessarily lead to declines in COVID-19 case counts without the implementation of extensive public health interventions.

## Introduction

Since December 2019, an increasing number of pneumonia cases caused by a novel coronavirus (COVID-19) have been identified in Wuhan, China (*1*). This new pathogen has exhibited high human-to-human transmissibility with approximately 43,112 confirmed cases and 1,018 deaths reported globally as of February 10, 2020.

On January 23, 2020, Wuhan - a city in China of 11 million residents - was forced to shut down both outbound and inbound traffic in an effort to contain the COVID-19 outbreak ahead of the Lunar New Year. However, it is estimated that more than five million people had already left the city before the lockdown (*3*), which has led to the rapid spread of COVID-19 within and beyond Wuhan.

In addition to population mobility and human-to-human contact, environmental factors can impact droplet transmission and survival of viruses (e.g., influenza) but have not yet been examined for this novel pathogen. Absolute humidity, defined as the water content in ambient air, has been found to be a strong environmental determinant of other viral transmissions (*4, 5*). For example, influenza viruses survive longer on surfaces or in droplets in cold and dry air - increasing the likelihood of subsequent transmission. Thus, it is key to understand the effects of environmental factors on the ongoing outbreak to support decision-making pertaining to disease control. Especially in locations where the risk of transmission may have been underestimated, such as in humid and warmer locations.

### Our contribution

We examine variability in absolute humidity and transmission of COVID-19 across provinces in China and other select locations. We show that the observed patterns of COVID-19 are not completely consistent with the hypothesis that high absolute humidity may limit the survival and transmission of this new virus.

## Data and Methods

### Epidemiological data

To conduct our analysis, we collected epidemiological data from the Johns Hopkins Center for Systems Science and System website (*6*). Incidence data were collected from various sources, including the World Health Organization (WHO); U.S. Centers for Disease Control and Prevention, China Center for Disease Control and Prevention CDC, European Centre for Disease Control and Prevention the Chinese National Health Center (NHC) as well as DXY, a Chinese website that aggregates NHC and local CCDC situation reports in near real-time. Daily cumulative confirmed incidence data were collected for each province in China from January 23, 2020 (i.e., the closure of Wuhan) to February 10, 2020. For comparison, we also obtained epidemiological data for affected countries including Thailand, Singapore, Japan, and South Korea, as well as other regions in China with important differences in data collection, including Hong Kong and Taiwan.

### Estimation of a proxy for the reproductive number

Based on the cumulative incidence data for each province, we estimated a proxy for the reproductive number *R* in a collection of 5, 6 and 7-day intervals (*7*). *R* is a measure of potential disease transmissibility defined as the average number of people a case infects before it recovers or dies. Our proxy for *R*, designated as *R*_*proxy*_, is a constant that maps cases occurring from time (*t*) to time (*t* + *d*) onto cases reported from time (*t* + *d*) to time (*t* + 2*d*); where *d* is an approximation of the serial interval (i.e., the number of days between successive cases in a chain of disease transmission). For multiple time points, *t*, we obtained values of *R*_*proxy*_(*t, d*) given by :

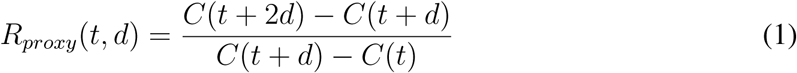

where the values of *d* range from [5 to 7], consistent with reported estimates of serial intervals (*7*).

Our measure is considered only a proxy for *R* because it does not use details of the (so far poorly defined) serial interval distribution, but instead, simply calculates the multiplicative increase in the number of incident cases over approximately one serial interval. Such proxies are at least approximately monotonically related to the true reproductive number and cross 1 when the true reproductive number crosses 1 (*8*). After computing these proxy values over a variety of subsequent moving time windows, a mean value was obtained and utilized as our estimated reproductive number *R* for each province. To minimize potential inclusion of imported cases in our analysis, *R* estimates were calculated after the closure of Wuhan on January 23rd, 2020.

### Weather data

We obtained monthly values of temperature and relative humidity data in January 2020 for each provincial capital in China from World Weather Online (*9*). We assumed that the majority of disease incidence for each province would occur or be reported in or near the capital due to increased population density in these metropolitan areas. We then calculated the “absolute humidity” from these two variables for each province using the following formula, which is an approximation of the Clausius Clapeyron equation (*10, 11*):

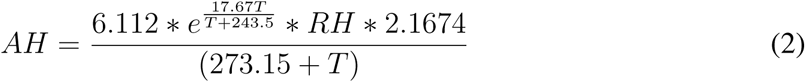

where *AH* is the absolute humidity, *T* is the temperature in degrees Celsius, *RH* is the relative humidity in percent (0-100), and *e* is the base of the natural *log*.

### Relationship between local exponential growth and environmental factors

We used two different interpolation approaches, a Loess regression and an exponential fit, to visually identify the relationship between absolute humidity and our proxy for the reproductive number (an indicator of the observed exponential growth rate) for each location. In order to identify the statistical relevance of the relationship between the local reproductive number of COVID-19 cases and environmental factors, we fitted a linear model using the logarithm of the local reproductive number *R*_*proxy*_ as our response variable. Absolute humidity and temperature were included as independent variables.

## Results

### Reproductive number proxy

Our estimates of *R*_*proxy*_, for each province within China and other countries appeared to be consistent across the range of serial intervals we analyzed, as shown by the vertical lines displayed in Figure 1 for each location. Most regions demonstrated *R*_*proxy*_ estimates well above 1, signaling the likely presence of local exponential growth.

**Figure 1:**
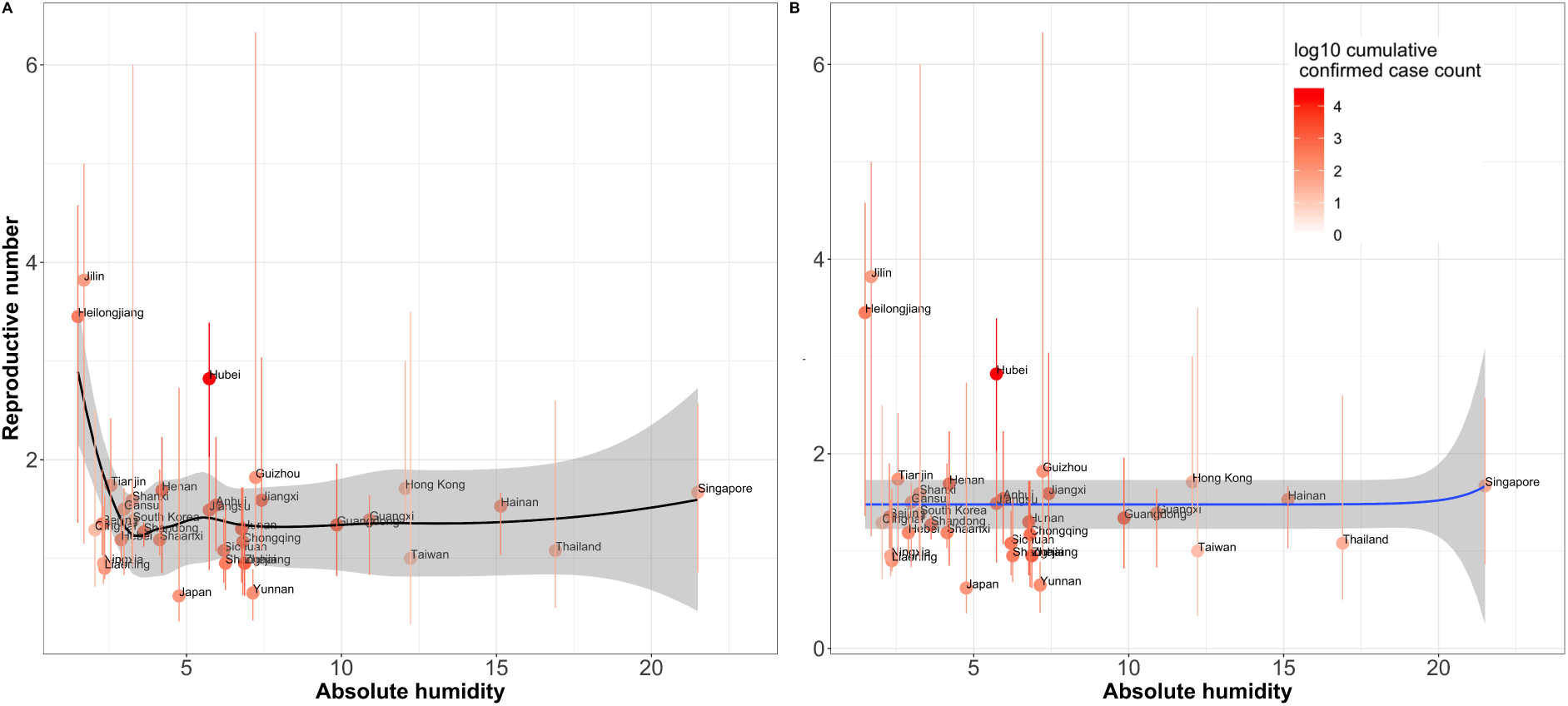
Estimated reproductive numbers *R*_*proxy*_ for COVID-19 plotted as a function of absolute humidity by province. 87% confidence intervals are displayed as vertical lines and were obtained from the collection of *R*_*proxy*_ calculated in subsequent time windows of length *d* for each location. Given the short time length of the current epidemic outbreak, an average of only 15 *R*_*proxy*_ values were calculated per location. Loess regression is shown to the left and exponential fit is shown to the right.

**Figure 2:**
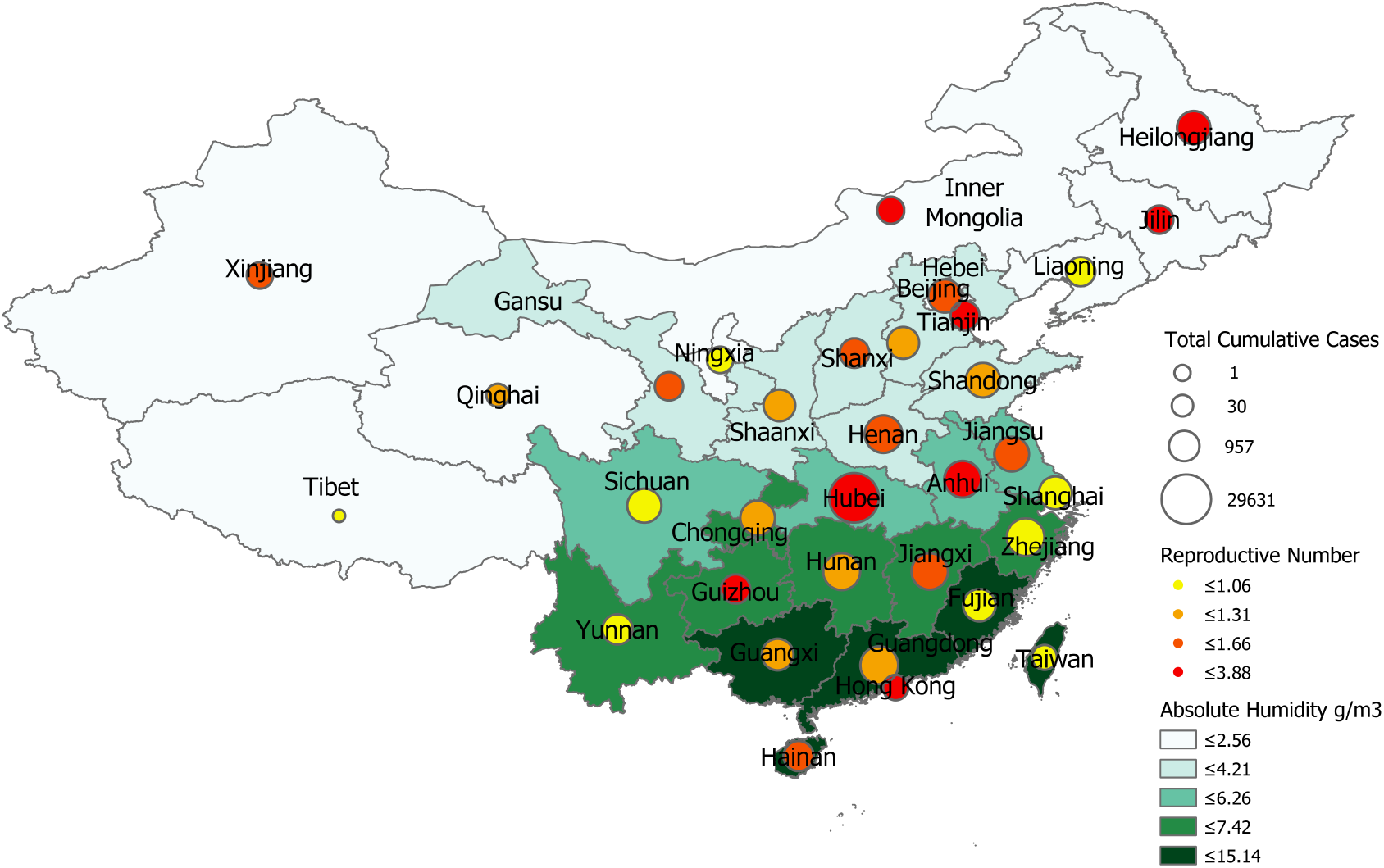
Absolute humidity in each provincial capital vs. COVID-19 *R*_*proxy*_ estimate (calculated after the closure of Wuhan city using data from 01/24/2020 to 02/10/2020). The size and color of each pin indicate cumulative cases per province and *R*_*proxy*_ range, respectively.

### Univariate relationship with absolute humidity

As shown by two separate data interpolation approaches in Figure 1, observed patterns of transmissibility as a function of absolute humidity were mixed. Specifically, panel A in Figure 1 (Loess regression) shows that not only dry and cold locations experience high values of *R*_*proxy*_ (as in influenza) but locations with high absolute humidity may also have higher values of *R*_*proxy*_, while the magnitude of its variability is small and all values are well above 1.

### Relationship with environmental factors

The regression model, demonstrates that both absolute humidity and temperature are associated with local exponential growth of COVID-19 across provinces in China and other affected countries (Table 1). Absolute humidity and temperature yielded a positive relationship and a slight negative relationship respectively.

**Table 1:** Relationship between local exponential growth, *log*(*R*_*proxy*_), and environmental factors (i.e., absolute humidity and temperature).

|  |  |
| --- | --- |
| Number of observations | 37 |
| F-statistic | 4.878 |
| P-value (F-statistic) | 0.01372 |
| R-squared | 0.223 |
| Adjusted R-squared | 0.1773 |

| Variable | Coefficient | Std Error | T-Statistic | P-value |
| --- | --- | --- | --- | --- |
| Intercept | $5.36 * 10^{-17}$ | 0.149 | 0.00 | 1.00 |
| Absolute Humidity | 0.761 | 0.370 | 2.054 | **0.048** |
| Temperature Mean | -1.050 | 0.370 | -2.836 | **0.008** |

### Limitations

Our estimates of the observed *R*_*proxy*_ across locations were calculated using available and likely incomplete reported case count data, with date of reporting, rather than date of onset, which adds noise to the estimation. In addition, the relatively short time length of the current outbreak, combined with imperfect daily reporting practices, make our results vulnerable to changes as more data becomes available. We have assumed that travel limitations and other containment interventions have been implemented consistently across provinces and have had similar impacts (thus population mixing and contact rates are assumed to be comparable), and have ignored the fact that different places may have different reporting practices. Further improvements could incorporate data augmentation techniques that may be able to produce historical time series with likely estimates of case counts based on onset of disease rather than reporting dates. This, along with more detailed estimates of the serial interval distribution, could yield more realistic estimates of *R*. Finally, further experimental work needs to be conducted to better understand the mechanisms of transmission of the COVID-19. Mechanistic understanding of transmission could lead to a coherent justification of our findings.

## Conclusion

Sustained transmission and rapid (exponential) growth of cases are possible over a range of humidity conditions ranging from cold and dry provinces in China, such as Jilin and Heilongjiang, to tropical locations, such as Guangxi and Singapore. Our results suggest that changes in weather alone (i.e., increase of temperature and humidity as spring and summer months arrive in the North Hemisphere) will not necessarily lead to declines in case counts without the implementation of extensive public health interventions. Further studies on the effects of absolute humidity and temperature on COVID-19 transmission are needed.

## Data Availability

All the data used for this study is publicly available as reported in the manuscript.

https://systemsjhuedu/research/public-health/ncov/

https://wwwworldweatheronlinecom/

## Notes

### Competing Interest Statement

The authors have declared no competing interest.

### Clinical Trial

NA

### Funding Statement

MS and CP are partially supported by the National Institute of General Medical Sciences of the National Institutes of Health under Award Number R01GM130668. The content is solely the responsibility of the authors and does not necessarily represent the official views of the National Institutes of Health.

## References and Notes

1. Zhu, N., et al., A novel coronavirus from patients with pneumonia in China, 2019. New England Journal of Medicine, 2020.

2. World Health Organization. Novel coronavirus (2019-nCoV). Available from: https://www.who.int/emergencies/diseases/novel-coronavirus-2019.

3. CGTN. Five million people left Wuhan before the lockdown, where did they go? Available from: https://news.cgtn.com/news/2020-01-27/5-million-people-left-Wuhan-before-the-lockdown-where-did-they-go–NACCu9wItW/index.html.

4. Barreca, A.I. and J.P. Shimshack, Absolute humidity, temperature, and influenza mortality: 30 years of county-level evidence from the United States. American journal of epidemiology, 2012. 176(suppl 7): p. S114–S122.

5. Shaman, J., E. Goldstein, and M. Lipsitch, Absolute humidity and pandemic versus epidemic influenza. American journal of epidemiology, 2011. 173(2): p. 127–135.

6. Johns Hopkins Center for Systems Science and System website https://systems.jhu.edu/research/public-health/ncov/

7. Li, Q., et al., Early Transmission Dynamics in Wuhan, China, of Novel Coronavirus-Infected Pneumonia. New England Journal of Medicine, 2020.

8. Wallinga, J. and Lipsitch, M., 2007. How generation intervals shape the relationship between growth rates and reproductive numbers. Proceedings of the Royal Society B: Biological Sciences, 274(1609), pp. 599–604.

9. World Weather online. https://www.worldweatheronline.com/

10. Iribarne, J.V. and W.L. Godson, Atmospheric thermodynamics. Vol. 6. 2012: Springer Science Business Media.

11. Bolton, D., 1980. The computation of equivalent potential temperature. Monthly weather review, 108(7), pp. 1046–1053.

